# Can co-designed educational interventions help consumers think critically about asking ChatGPT health questions? Results from a randomised-controlled trial

**DOI:** 10.1101/2025.04.28.25326600

**Authors:** Julie Ayre, Melody Taba, Brooke Nickel, Geoffrey Edlund, Trang Vu, Julia Yan, Lorna Butters, Ivan C.K. Ma, Kirsten J. McCaffery

**Author notes:** Corresponding author: Dr Julie Ayre. Australia and New Zealand Clinical Trials registration: ACTRN 12624000641594.

## Abstract

**Background:** Generative artificial intelligence (AI) tools have clear potential health benefits for individuals (e.g. simplifying information) and risks (e.g. inaccurate information).

**Objective:** To evaluate two brief co-designed health literacy interventions to help people critically reflect on health questions they ask ChatGPT.

**Design:** Three arm parallel-group randomised controlled trial

**Participants:** Australian adults without university education who had used ChatGPT in the past 6 months, and recruited via online social research panel.

**Interventions:** (1) animation intervention, (2) image-based intervention, or (3) control.

**Main measures:** The primary outcomes were intention to ask ChatGPT a question in ‘lower risk’ and ‘higher risk’ scenarios, where higher risk scenarios would typically require clinical interpretation. Secondary outcomes were ChatGPT knowledge, trust in ChatGPT’s responses, and intervention acceptability.

**Key results:** Of the 619 participants, 592 were included in the analysis sample. Average age was 47.0 years (SD=16.4), 42.6% identified as man or male, and 17.4% had limited/marginal health literacy. Participants in the animation group (n=191) reported lower intention to use ChatGPT for higher risk scenarios (M=2.42/5, 95%CI: 2.27 to 2.56) compared to those in the images group (n=203, M=2.69/5, 95%CI: 2.54 to 2.83, p=0.010). Participants in both intervention groups reported lower intentions to use ChatGPT for higher risk scenarios compared to control (n=205, M=3.12/5, 95%CI: 2.98 to 3.27, p<0.001). There was no effect of intervention group on intention to use ChatGPT for lower risk scenarios (p=0.800). Participants in the intervention groups had higher knowledge of ChatGPT (p<0.001) and reported lower trust (p<0.001), compared to those in the control group.

**Conclusions:** Brief health literacy interventions may help improve knowledge of ChatGPT and reduce intentions to ask riskier health questions. This study represents an initial step towards addressing AI health literacy and highlights the kinds of health literacy skills that can support people to navigate AI tools safely.

## Introduction

Generative artificial intelligence (AI) tools such as ChatGPT allow users to ask any question and immediately receive a plausible and highly tailored response. The uptake of generative AI tools has increased steadily over time and there is now a sizable portion of the population using them. For example, an estimated 1 in 10 Australians reported using ChatGPT to ask health questions in the first half of 2024, with a further 3 in 10 considering asking it a health question in the next 6 months.^1^ Since then, Google has added an ‘AI overview’ to search results in over 100 countries, bringing generative AI to one of the most widely used internet tools.^2^ Several commentaries have expressed a hesitant optimism about the increasingly prominent role of generative AI in health communication, highlighting the potential opportunities and risks.^3-5^

On the one hand, generative AI tools have the potential to support informed health decision-making by improving the accessibility of health information. Several studies have now shown that free tools such as ChatGPT can significantly improve the readability of health information,^6,7^ with newer models such as GPT-4 delivering content that meets grade reading level targets whilst maintaining a high level of accuracy ^8^. This is an important capability as research consistently reports that online health information is overly complex.^9^

However, research shows that the accuracy of generative AI tools can vary depending on several factors. For example, some reviews have suggested that accuracy tends to be higher for general or factual health questions, and lower for questions that require critical thinking, situational awareness (context of personal/social situation and medical history), awareness of regional guidelines, specific treatment recommendations, and questions relating to rare diseases.^6,10-14^

The accuracy can also vary according to the types of prompts and evaluation metrics used.^11^ Errors and hallucinations (made up facts) do occur and pose a genuine risk to patient safety, such as errors in medication instructions.^15^ These issues are particularly pertinent for people using generic tools such as ChatGPT, as these models have not been specifically trained to answer health queries, nor do they draw on a specialised medical knowledge base.

As generative AI tools become more widely used, the public will need to acquire the knowledge and skills to help them take advantage of these tools whilst also avoiding the potential risks. It is critical that efforts to improve knowledge and skills are accessible to people with limited health literacy and who do not speak the dominant language of the country they live in, as these groups are more likely to use these tools for health tasks.^1^ and stand to benefit substantially from access to free, tailored, understandable, and translated health content. Although frameworks and interventions for digital health literacy are increasingly common,^16,17^ none of these to date have explicitly considered the skills needed to navigate generative AI tools safely.^18^

This study sought to bridge this gap in the research by co-designing and evaluating two brief health literacy interventions. These interventions aim to equip users with foundational knowledge about ChatGPT and the skills to reflect on the potential risks and benefits of using it to answer their health questions. We hypothesised that animation and imaged-based educational resources would improve AI health literacy skills relative to controls. Given that previous health literacy intervention studies have shown that short animations are often more effective than text-based resources,^19^ we also hypothesised that the animation would be more effective than the image-based resource.

## Methods

### Study design

This was a co-designed three arm parallel-group randomised controlled trial study design with participants randomly allocated to intervention group in the ratio 1:1:1 (Figure 1). This trial was prospectively registered with the Australian New Zealand Clinical Trials Registry (ACTRN 12624000641594) and approved by the University of Sydney Human Research Ethics Committee (2024/HE000247). CONSORT checklist is available in Appendix A. This online study was available through the Qualtrics platform.

**Figure 1.**
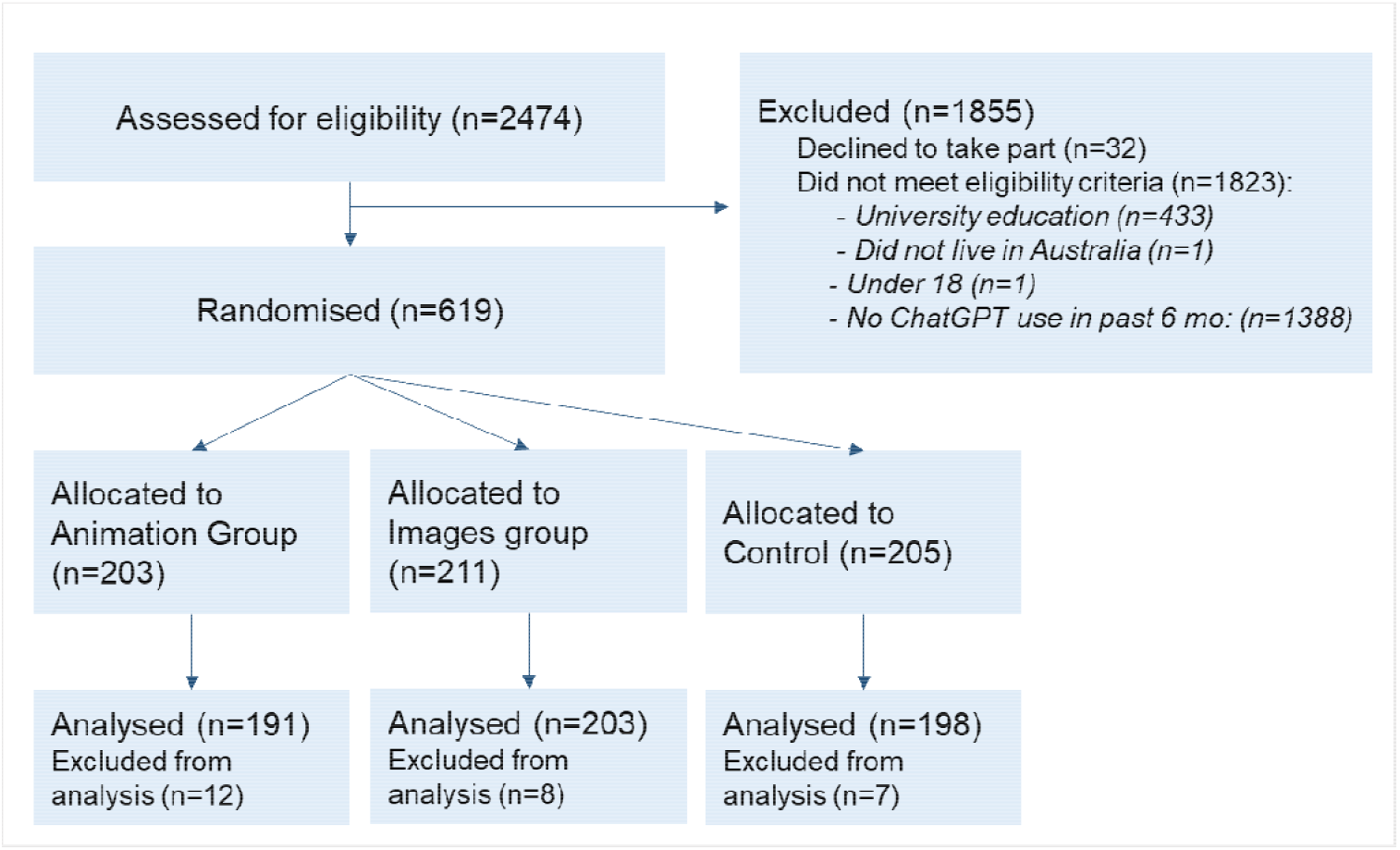
Participant flow and study design.

### Community involvement

The research question was initially proposed by a community member after reflecting on a previous ChatGPT study that they had contributed to.^7^ The current study was supported by a panel of 6 community members with diverse experiences (age, gender, cultural background, metropolitan vs rural, and familiarity with ChatGPT). Members met online across 4 meetings between November 2023 and May 2024 (Table S1). Community involvement was integral to refining key messages (Table S2), co-development of resources, and designing the study including purpose-built outcome measures to evaluate knowledge and skills. A fifth meeting sought to integrate community perspectives into the interpretation of the results.

### Participants

Eligible participants were adults (18+ years) living in Australia who did not have a university level of education, and who reported using ChatGPT in the past 6 months. Participants were recruited between 20 November and 16 December 2025, through PureProfile, which manages a member panel of over 550,000 Australian adults.

### Intervention groups

The key messages that were conveyed in the intervention resources were developed between November 2023 to May 2024, and reflect the capabilities of ChatGPT during this period (Table S2). These key messages continued to be of relevance during the recruitment period. Both intervention (images and animation) covered the same content.

### Animation intervention group

Participants viewed an animation (3min 40s) that included a voiceover and closed captions. The script was written at a Grade 9 reading level and contained 7.3% complex language and 1 passive voice, as assessed by the Sydney Health Literacy Lab Health Literacy Editor.^20^ Participants had the ability to pause, replay, and change video speed.

### Image intervention group

Participants viewed a series of 7 images embedded within an image carousel, similar to what appears on the social media platform Instagram. In addition to the key messages shown in Table S2, the images showed examples of questions that you might try or avoid asking ChatGPT. The text was written at a Grade 7.8 reading level and contained 7.3% complex language, with no passive voice. Participants were able to move backwards and forwards through the images.

### Control

Participants viewed an infographic on healthy eating that had been developed by the Australian Government.^21^ This infographic was selected as a control as it was a neutral health stimulus that did not contain any content related to ChatGPT or online health information seeking.

### Procedure

Piloting took part between 5 July and 7 November 2025, and ensured that recruitment methods were feasible and to refine the phrasing of the primary outcome items to ensure they were clear.

Potential participants were invited to take part via the PureProfile portal, with links to the survey hosted on Qualtrics. After consenting, participants completed demographic items, a health literacy screening item^22^ and a measure of digital health literacy (eHeals).^23^ Participants were asked how much they trusted ChatGPT, how often they had used ChatGPT in the past 6 months, and what kinds of health questions they had asked ChatGPT (Appendix B).

Participants were then randomised to an intervention group and asked to view the materials. Participants in the animation group were required to remain on the page for the length of the video (3 minutes 40 seconds). Participants in the image and control groups were required to remain on the page for 60 seconds. All participants then completed primary and secondary outcomes, and those in the intervention groups completed questions about acceptability of the intervention. Participants who completed the survey received reimbursement for their participation.

### Outcomes

#### Primary outcome

The primary outcomes were intentions to use ChatGPT for lower and higher risk health questions. These were based on responses to 7 co-designed, scenario-based items that asked participants to indicate their intentions to ask different health questions to ChatGPT (5-point Likert scale: definitely avoid to definitely try). Scenarios for each item are shown in Appendix B. Items 1, 2, 4 and 6 were considered lower risk as they focused on scenarios about general health knowledge. Items 3, 5 and 7 were considered higher risk as they focused on scenarios asking for personalised advice about treatment, pathology and diagnosis, that would usually require clinical interpretation. These scores assessed intention to use ChatGPT for higher risk scenarios. Though a measure of intention, scores for this outcome reflect participants’ ability to apply their ChatGPT knowledge and skills to novel hypothetical health scenarios.

#### Secondary outcomes

Participants also completed a 5-item assessment of ChatGPT knowledge (Appendix B). These items mirrored key messages in the intervention content and were marked as correct or incorrect. Participants repeated the baseline assessment of trust in ChatGPT after viewing the interventions.

#### Acceptability outcomes

Those allocated to the two intervention groups completed items relating to intervention acceptability (Appendix B). Perceived effectiveness was assessed via 6 descriptors (worth-remembering, attention-grabbing, powerful, informative, meaningful and convincing), with responses recorded on a 5-point Likert scale.^24^ Intention to share the intervention on social media was collected using a single item adapted from another digital health literacy intervention study (7-point Likert scale).^25^ Seven items assessed perceived usefulness of the intervention content, with responses rated using a 5 point-Likert scale. Items asked about novelty of the intervention, its impact, future intentions to act on advice in the intervention, and confidence in the intervention content. This was adapted from the same digital health literacy intervention study.^25^ All scales ranged from strongly disagree to strongly agree.

#### Analysis

Planned contrasts between the two intervention arms and control arm were implemented in regression models (i.e. interventions vs control; animation vs images). Exploratory analyses examined the influence of health literacy, digital literacy, gender, age, and trust in ChatGPT, by including interaction terms within the regression models. These were analysed as binary categories. Categories for health literacy reflect the cut-offs suggested by Wallace et al. ^22^ (i.e. for limited/marginal and adequate). Scores below the midpoint of the eHeals scale were considered ‘low’ digital health literacy. For trust in ChatGPT, scores of ‘somewhat’ or below were considered ‘low.’ Two age categories (18 to 44 and 45+) were used as these age groups observed varied use of ChatGPT for health in a previous study.^1^ Outcomes related to acceptability were analysed descriptively.

In line with existing fraud mitigation strategies, participants were excluded from analyses if they completed survey questions in a short timeframe (i.e. less than 2 minutes for survey questions estimated to take 9 minutes) or failed to meet built-in Qualtrics quality checks e.g. ReCAPTCHA scores.^26^ All participants who met eligibility and fraud mitigation criteria passed the commitment check.

#### Sample size

The total sample size required was 567. This was based on an expected small effect size of 0.15 for the primary outcome. The number of participants required per arm (3 arms), to provide 90% power at error type I (alpha =0.05) was 189 (this is a balanced design).

## Results

619 participants were randomised to an intervention group, and the analysis sample of 592 participants excluded 27 who did not meet fraud mitigation criteria (Figure 1). Descriptive statistics for the analysis sample are shown in Table 1. Two hundred and fifty-two participants (42.6%) identified as man or male, average age was 47.0 years (SD=16.4), and 17.4% had limited/marginal health literacy using the screening item (n=103). On average, participants scored above the midpoint of the eHealth literacy scale (M=3.7/5, SD=0.7). All participants had used ChatGPT in the past 6 months, and two fifths had not asked the tool a health question during this period (n=245, 41.4%). A similar proportion reported asking health questions a few times during this period (39.5%, n=234). Participants reported on average ‘somewhat’ trusting ChatGPT’s advice (M=3.00/5, SD=0.91). The most common health questions that they considered asking ChatGPT related to learning about a health condition (33.8%; n=200), finding out what symptoms meant (30.4%, n=180), and understanding medical terms (23.6%, n=140).

**Table 1.**
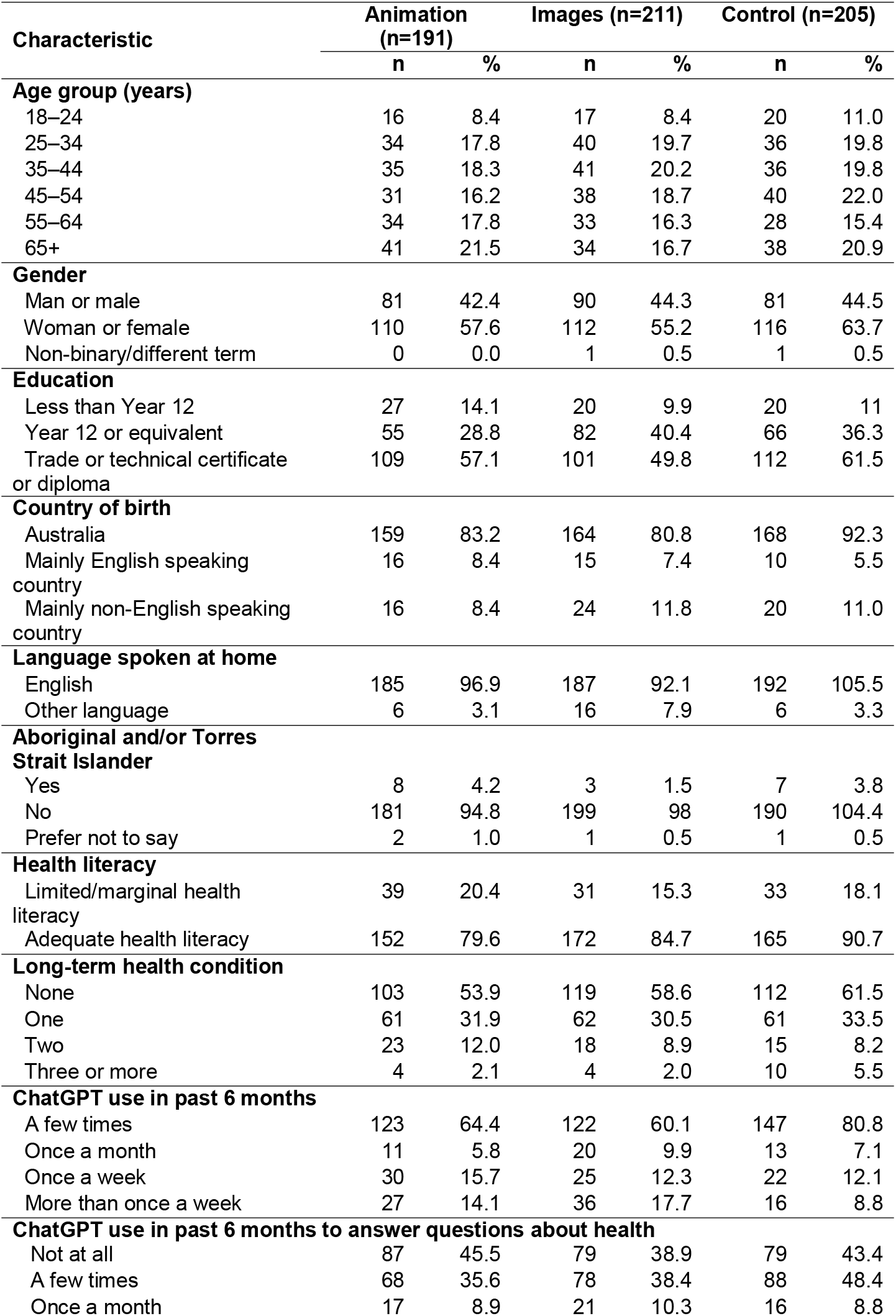
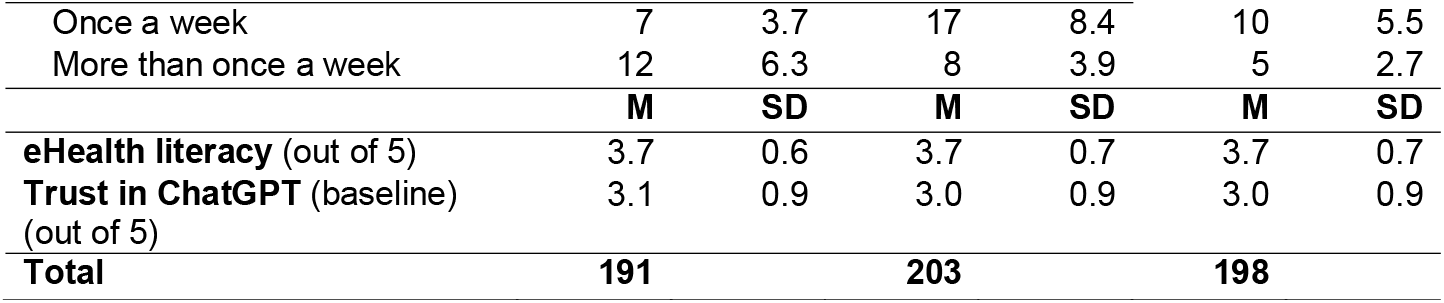
Participant characteristics, N=592.

| Characteristic | Animation (n=191) |  | Images (n=211) |  | Control (n=205) |  |
| --- | --- | --- | --- | --- | --- | --- |
|  | n | % | n | % | n | % |
| **Age group (years)** |  |  |  |  |  |  |
| 18–24 | 16 | 8.4 | 17 | 8.4 | 20 | 11.0 |
| 25–34 | 34 | 17.8 | 40 | 19.7 | 36 | 19.8 |
| 35–44 | 35 | 18.3 | 41 | 20.2 | 36 | 19.8 |
| 45–54 | 31 | 16.2 | 38 | 18.7 | 40 | 22.0 |
| 55–64 | 34 | 17.8 | 33 | 16.3 | 28 | 15.4 |
| 65+ | 41 | 21.5 | 34 | 16.7 | 38 | 20.9 |
| **Gender** |  |  |  |  |  |  |
| Man or male | 81 | 42.4 | 90 | 44.3 | 81 | 44.5 |
| Woman or female | 110 | 57.6 | 112 | 55.2 | 116 | 63.7 |
| Non-binary/different term | 0 | 0.0 | 1 | 0.5 | 1 | 0.5 |
| **Education** |  |  |  |  |  |  |
| Less than Year 12 | 27 | 14.1 | 20 | 9.9 | 20 | 11 |
| Year 12 or equivalent | 55 | 28.8 | 82 | 40.4 | 66 | 36.3 |
| Trade or technical certificate or diploma | 109 | 57.1 | 101 | 49.8 | 112 | 61.5 |
| **Country of birth** |  |  |  |  |  |  |
| Australia | 159 | 83.2 | 164 | 80.8 | 168 | 92.3 |
| Mainly English speaking country | 16 | 8.4 | 15 | 7.4 | 10 | 5.5 |
| Mainly non-English speaking country | 16 | 8.4 | 24 | 11.8 | 20 | 11.0 |
| **Language spoken at home** |  |  |  |  |  |  |
| English | 185 | 96.9 | 187 | 92.1 | 192 | 105.5 |
| Other language | 6 | 3.1 | 16 | 7.9 | 6 | 3.3 |
| **Aboriginal and/or Torres Strait Islander** |  |  |  |  |  |  |
| Yes | 8 | 4.2 | 3 | 1.5 | 7 | 3.8 |
| No | 181 | 94.8 | 199 | 98 | 190 | 104.4 |
| Prefer not to say | 2 | 1.0 | 1 | 0.5 | 1 | 0.5 |
| **Health literacy** |  |  |  |  |  |  |
| Limited/marginal health literacy | 39 | 20.4 | 31 | 15.3 | 33 | 18.1 |
| Adequate health literacy | 152 | 79.6 | 172 | 84.7 | 165 | 90.7 |
| **Long-term health condition** |  |  |  |  |  |  |
| None | 103 | 53.9 | 119 | 58.6 | 112 | 61.5 |
| One | 61 | 31.9 | 62 | 30.5 | 61 | 33.5 |
| Two | 23 | 12.0 | 18 | 8.9 | 15 | 8.2 |
| Three or more | 4 | 2.1 | 4 | 2.0 | 10 | 5.5 |
| **ChatGPT use in past 6 months** |  |  |  |  |  |  |
| A few times | 123 | 64.4 | 122 | 60.1 | 147 | 80.8 |
| Once a month | 11 | 5.8 | 20 | 9.9 | 13 | 7.1 |
| Once a week | 30 | 15.7 | 25 | 12.3 | 22 | 12.1 |
| More than once a week | 27 | 14.1 | 36 | 17.7 | 16 | 8.8 |
| **ChatGPT use in past 6 months to answer questions about health** |  |  |  |  |  |  |
| Not at all | 87 | 45.5 | 79 | 38.9 | 79 | 43.4 |
| A few times | 68 | 35.6 | 78 | 38.4 | 88 | 48.4 |
| Once a month | 17 | 8.9 | 21 | 10.3 | 16 | 8.8 |
| Once a week | 7 | 3.7 | 17 | 8.4 | 10 | 5.5 |
| More than once a week | 12 | 6.3 | 8 | 3.9 | 5 | 2.7 |
|  | **M** | **SD** | **M** | **SD** | **M** | **SD** |
| **eHealth literacy** (out of 5) | 3.7 | 0.6 | 3.7 | 0.7 | 3.7 | 0.7 |
| **Trust in ChatGPT** (baseline)<br>(out of 5) | 3.1 | 0.9 | 3.0 | 0.9 | 3.0 | 0.9 |
| **Total** | **191** |  | **203** |  | **198** |  |

### Evaluation of the educational resources

#### Primary outcome

There was no effect of intervention group on intention to use ChatGPT for lower risk scenarios (F_(1,589)_=0.22, p=0.800; Table 2, Figure 2). Participants in the animation group reported lower intention to use ChatGPT for higher risk scenarios (M=2.42, SE=0.07, 95%CI: 2.27 to 2.56) compared to those in the images group (M=2.69, SE=0.07, 95%CI: 2.54 to 2.83, p=0.010). Participants in both intervention groups reported lower intentions to use ChatGPT for higher risk scenarios compared to control (M=3.12, SE=0.07, 95%CI: 2.98 to 3.27, p<0.001).

**Table 2.** Intention to use ChatGPT for health questions by intervention group*.

| ChatGPT health scenario | Animation |  | Images |  | Control |  |
| --- | --- | --- | --- | --- | --- | --- |
|  | M | SD | M | SD | M | SD |
| Learning about a health condition | 4.23 | 0.89 | 4.11 | 1.08 | 4.06 | 1.06 |
| Learning about treatment for a health condition | 3.94 | 0.93 | 4.03 | 1.00 | 4.08 | 0.92 |
| Asking for advice about a specific treatment | 2.24 | 1.19 | 2.50 | 1.22 | 2.98 | 1.19 |
| Asking about what a blood test measures | 3.96 | 1.03 | 3.92 | 1.08 | 3.84 | 0.98 |
| Asking to interpret a blood test result | 2.62 | 1.23 | 2.80 | 1.28 | 3.26 | 1.18 |
| Asking to simplify reputable information | 3.63 | 1.06 | 3.53 | 1.12 | 3.60 | 0.99 |
| Asking about whether to see a doctor based on symptoms | 2.39 | 1.29 | 2.76 | 1.40 | 3.13 | 1.22 |
| **Lower risk scenarios** | **3.94** | **0.72** | **3.90** | **0.84** | **3.89** | **0.81** |
| **Higher risk scenarios** | **2.42** | **1.02** | **2.69** | **1.09** | **3.12** | **0.99** |

\*Higher scores indicate higher intention of using ChatGPT. A score of 1 refers to 'definitely avoid' and a score of 5 refers to 'definitely try' to use ChatGPT to answer the health question in the scenario(s). Scenarios categorised as higher risk are shown in grey.

**Figure 2.**
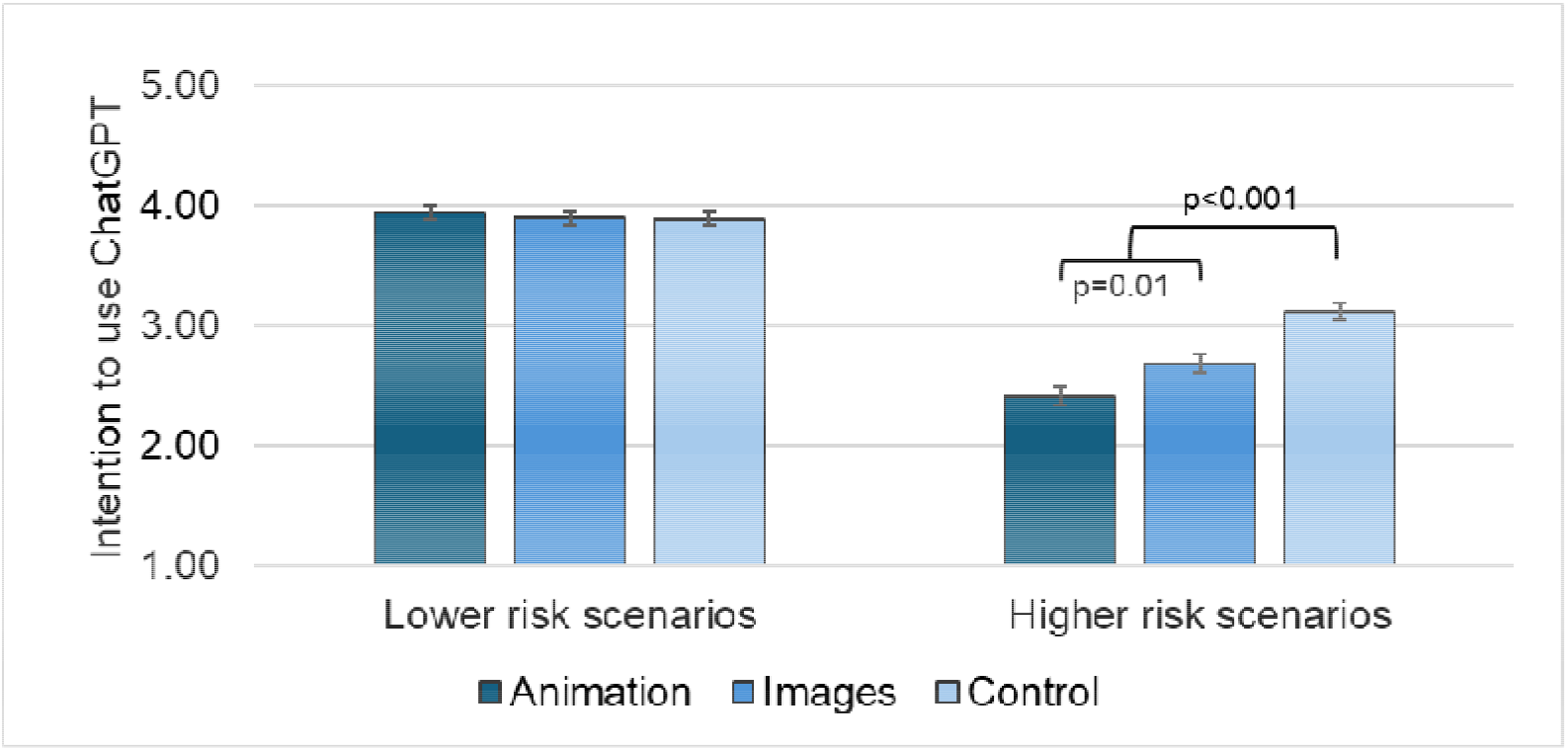
Intention to use ChatGPT, by intervention group and scenario type. *Note*: Error bars indicate ±1 SE. Higher scores indicate greater intention (possible range 1 to 5).

#### Secondary outcomes

Participants in the animation group obtained higher ChatGPT knowledge scores (M=4.13, SE=0.09, 95%CI: 3.94 to 4.31) than those in the image group (M=3.7, SE=0.09, 95%CI: 3.54 to 3.89, p=0.001; Figure 3, Tables S3 and S4). Participants in both intervention groups obtained higher knowledge scores than those in the control group (M=3.14, SE=0.09, 95%CI: 2.96 to 3.31, p<0.001).

**Figure 3.**
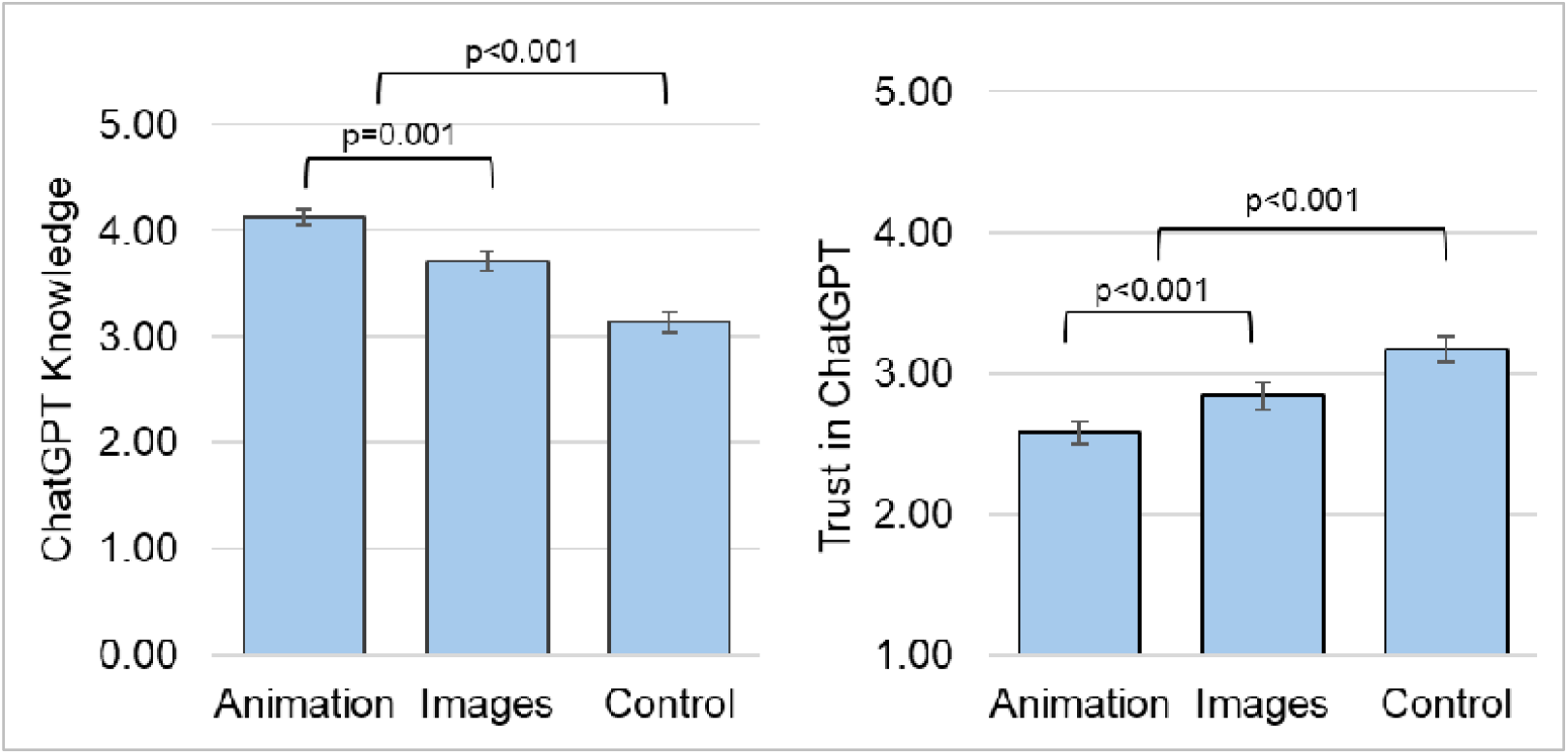
ChatGPT knowledge and trust, by intervention group. *Note*: Possible knowledge score range is 0 to 5. Possible trust score range is 1 to 5, where higher scores indicate greater trust. This figure shows estimated marginal means for trust at follow-up, controlling for trust at baseline.

Controlling for baseline trust in ChatGPT, participants in the animation group reported lower trust in ChatGPT (M=2.58, SE=0.05, 95%CI: 2.48 to 2.68) than those in the image group (M=2.84, SE=0.05, 95%CI: 2.74 to 2.94, p<0.001; Figure 3, Table S3). Participants in both intervention groups reported lower trust in ChatGPT than those in the control group (M=3.17, SE=0.05, 95%CI: 3.07 to 3.27, p<0.001).

#### Exploratory analyses

##### Lower risk scenarios

Across all intervention groups, participants with limited/marginal health literacy, low digital health literacy, and low (baseline) trust in ChatGPT on average had lower intentions to use ChatGPT in lower risk scenarios (Tables S5 and S6). Younger participants (aged 18 to 44 years) in the control group reported lower intentions to use ChatGPT for lower risk scenarios (M=3.65, 95% CI: 3.49 to 3.81) compared to older participants (45 years or more) (M=4.10, 95% CI: 3.95 to 4.25; Tables S5 and S7; Figure S1). No other effects were observed for gender and age group.

##### Higher risk scenarios

For higher risk scenarios, we did not observe any significant main effects or interaction effects for health literacy, digital health literacy, gender, or age group (Table S5). People with low baseline trust in ChatGPT were less inclined to use the tool for higher risk scenarios (M=2.60, 95%CI: 2.51 to 2.70) compared to participants with high trust in ChatGPT (M=3.08, 95%CI: 2.93 to 3.23, p<0.001). This observation was more marked for participants in the control group (Tables S5 and S6, p=0.007; Figure S2).

#### Intervention acceptability

Participants rated the perceived effectiveness of the animation as 3.87/5 (SD=0.67) perceived effectiveness of the images as 3.56/5 (SD=0.74). Participants in both intervention groups rated their intentions to share the resource with friends or family above the midpoint of the scale (Animation: M=4.77/7, SD=1.4; Images: M=4.45, SD=1.51). Participants rated both the animation and the images as useful (Animation: M=4.07/5, SD=0.61; Images: M=3.93/5, SD=0.62).

## Discussion

This study provides world-first evidence that co-designed health literacy interventions can help people develop skills to critically appraise which kinds of health questions are riskier to ask ChatGPT. In line with the intervention content, people allocated to the animation and images intervention groups had lower intentions to ask ChatGPT health questions that are riskier, such as asking for personalised treatment advice, interpreting test results, or symptom diagnosis. Their intentions to ask lower risk health questions were similar to those of people in the control group. People allocated to the intervention groups also demonstrated significantly higher knowledge of ChatGPT and its limitations, and lower trust in the tool. These effects were more pronounced for people who viewed the animation compared to the images.

Reliance on ChatGPT and other generative AI tools as a source of health information is a contentious issue.^3,4^ Whilst there are clear benefits such as their ability to simplify health information,^8^ this must be balanced against the potential safety risks.^11^ The current study demonstrates that brief online interventions can develop AI health literacy skills and improve community use of this new technology. However, the exact nature of AI health literacy skills is likely to change as the technology advances. For example, ChatGPT can now integrate real information sources into its answers.^27^ This feature overcomes some of the tool’s key limitations, though the quality of the information sources remains a concern and still requires users to have critical appraisal skills.^28^

This study has several strengths and limitations. As described above, the project was initiated by a consumer and co-designed. We recruited a sample of participants who did not have an undergraduate level of education. This group is more likely to have limited/marginal health literacy and stand to benefit from interventions that incorporate health literacy principles.^29^

The study also has several limitations. There were no existing appropriate validated assessments of AI health literacy or ChatGPT knowledge that were available at the time of the study. Although we developed our own purpose-designed instrument, its psychometric properties have not been evaluated. We also acknowledge that the interventions were tested in a controlled setting, requiring participants to be exposed to the intervention for a specified time period. It is not clear how effective these interventions would be in real-life settings. For example, if implemented directly through social media, they may not compete effectively with the vast amount of ChatGPT content that rapidly adapts to incorporate trends in user-generated content. Strategies such as shorter videos, humour or memes, and content creation by influencers may increase engagement and intention to share the content,^30^ or alternatively the videos could be delivered in settings where longer content is more appropriate e.g. primary care clinics.

Future work could consider enhancing the content in several ways. The content could be updated for changes to ChatGPT technology and a broader skillset e.g. strategies to support preparation for medical consultations, adapted for other AI tools such as Gemini or Co-Pilot; and focus on cultivating broader skills to help people decide which AI tools to use. The content could also be adapted for people in culturally and linguistically diverse communities and translated into other languages, as we know from previous research that these group are more likely to ask ChatGPT health questions.^1^

Our findings demonstrate that brief health literacy interventions can help improve knowledge of ChatGPT and reduce intentions to ask riskier health questions. The interventions specifically targeted ChatGPT use; as such this study represents an initial step towards addressing AI health literacy in the community. It highlights the kinds of health literacy skills that can support people to navigate AI tools more safely. With the rapid uptake of AI tools such as ChatGPT in the community, it is crucial that health literacy and public health research keeps pace. This means developing equitable and inclusive resources to equip the community with skills they need to suit the changing digital environment.

## Supporting information

Appendix A CONSORT

Appendix B Survey

## Acknowledgement

We would like to acknowledge the contribution of Atria Rezwan, who suggested the idea for resources that help people more skilfully use ChatGPT to find answers to health questions. Julie Ayre, Kirsten McCaffery and Brooke Nickel are supported by National Health and Medical Research Council fellowships (APP2017278, APP2016719, APP1194108). These findings have been uploaded as a preprint (doi:). They have not been presented at any conferences.

## Declaration statement

The SHeLL Editor is a research tool owned by the University of Sydney. It is sublicensed to Health Literacy Solutions PTY Ltd to enable wider public use. Julie Ayre and Kirsten McCaffery are co-directors of Health Literacy Solutions PTY Ltd.

## Data availability statement

The datasets analysed during the current study available from the corresponding author on reasonable request

