## Appendix B Survey for "Can co-designed educational interventions help consumers think critically about asking ChatGPT health questions? Results from a randomised-controlled trial"

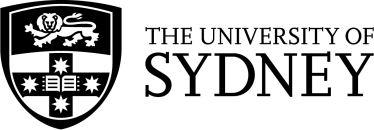

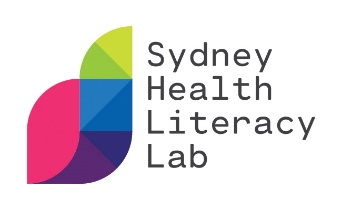

***Evaluating social media interventions
 to support safe use of ChatGPT for health***

1. **What is this study about?**

You may have heard about ChatGPT in the news or on social media. ChatGPT is a publicly available AI chatbot that is free, easy to use and gives quick, human-like responses to user questions and requests. We want to test two interventions to see if they help people use ChatGPT more safely when asking health questions.

2. **Who can take part in the study?**

You can take part of the study if you live in Australia and are over 18 years of age.

3. **What will I be asked to do?**

We will ask you to complete an online survey that will ask questions about you, for example, your age, gender and education. We will also ask you to complete some questions about the intervention and how you feel about asking ChatGPT about your health.

We expect this will take you about **15** **minutes**.

4. **Do I have to take part? Can I change my mind once I’ve started?**

Being in this study is completely voluntary and you do not have to take part. If you decide to take part, you can withdraw any time before you submit the survey. However, once your responses are submitted, they cannot be withdrawn. This is because they are anonymous, and we will not be able to tell which one yours is.

Your decision will not affect your current or future relationship with the researchers or anyone else at The University of Sydney.

To read more information about this part of the study, please download the Participant Information Statement <link to PDF version of PIS>. This study has received Ethics approval from University of Sydney Human Research Ethics Committee (Project No. 2024/255).

**I confirm that I have read the Participant Information Statement and consent to take part in this research project as described.**

| **Yes** | **No** |
| --- | --- |

[If ‘yes,’ proceed to survey question on next page, if ‘no,’ survey ends and no data is collected.]

***[Demographic questions]***

Thank you for taking part in our study. First, we have a few questions about you:

In the last 6 months how often have you used ChatGPT?

| Not at all | A few times | Once a month | Once a week | More than once a week |
| --- | --- | --- | --- | --- |

[Screen out participants who have not used ChatGPT at all]

In the last 6 months how often have you used ChatGPT to answer questions about health?

| Not at all | A few times | Once a month | Once a week | More than once a week |
| --- | --- | --- | --- | --- |

How old are you?

_____________________________________________________________________

[Screen out participants are less than 18 years]

Do you live in Australia?

- Yes
- No [direct to end of survey]

[Screen out participants who do not live in Australia]

What is the highest level of education you have **completed**?

- Less than Year 12 or equivalent
- Completed Year 12 or equivalent
- Trade or technical certificate or diploma
- University degree
- Postgraduate/higher degree

[Screen out participants who have respond University degree or postgraduate/higher degree]

What is your gender identity?

- Female/ Woman
- Male/ Man
- Non-binary
- I use another term (please specify)

______________________________________________________________

- Prefer not to say

2) In which country were you born?
   - Australia
   - Other (please tell us) ________

[If ‘other’ answered in previous question] In what year did you move to Australia?

__________________________________________________________________

What language do you speak at home?

- English
- Other (please tell us)

Are you of Aboriginal or Torres Strait Islander origin?

- Yes
- No
- Prefer not to say

Which, if any, of the following long-standing health conditions do you have (including age-related conditions)? (select all that apply)

| Deafness or severe hearing impairment | A neurological condition (e.g. Alzheimer’s, Parkinson’s) |
| --- | --- |
| Blindness or severe vision impairment | None of these |
| A longstanding illness (e.g. cancer, HIV, diabetes, chronic heart disease) |  |
| A longstanding physical condition (e.g. arthritis, spinal injury, multiple sclerosis) |  |
| An intellectual disability |  |
| A mental health condition (e.g. depression) |  |

[health literacy]

The next few questions ask you about your experience understanding health information.*****

a) If you need to go to the doctor, clinic, or hospital, how confident are you filling out the medical forms by yourself?

| Not at all confident | A little confident | Somewhat confident | Quite confident | Extremely confident |
| --- | --- | --- | --- | --- |

b) How often do you have someone (family member or staff at the clinic or hospital) help you to read health or medical information forms?

| Always | Often | Sometimes | Occasionally | Never |
| --- | --- | --- | --- | --- |

c) How often do you have problems learning about your health because of trouble understanding written health information?

| Always | Often | Sometimes | Occasionally | Never |
| --- | --- | --- | --- | --- |

d) How often do you have trouble understanding what your doctor, nurse, or pharmacist tells you about your health or about health information?

| Always | Often | Sometimes | Occasionally | Never |
| --- | --- | --- | --- | --- |

e) How often do you have trouble remembering instructions from the doctor, nurse, or pharmacist after you get home?

| Always | Often | Sometimes | Occasionally | Never |
| --- | --- | --- | --- | --- |

*The first item (a) is a validated single-item heath literacy screener. This was used in the analysis, rather than the full five item scale.

[Digital health literacy]

The next questions will ask about using the internet for health information. For each statement, tell me which response best reflects your experience *right now*.	(5-point scale strongly disagree to strongly agree)

1) I know **what** health resources are available on the Internet
2) I know **where** to find helpful health resources on the Internet
3) I know **how** to find helpful health resources on the Internet
4) I know **how to use** the Internet to answer my questions about health
5) I know how to use **the health information** I find on the Internet to help me
6) I have the skills I need to **evaluate** the health resources I find on the Internet
7) I can tell **high quality** health resources from **low quality** health resources on the Internet
8) I feel **confident** in using information from the Internet to make health decisions

In the last 6 months have you used ChatGPT to... (select all that apply)

- Find out what my symptoms mean (or the symptoms of someone I know)
- Find out what to do about a specific health issue that I or someone I know has
- Learn about a specific health condition
- Learn about healthy lifestyles
- Help create a plan to improve my health (or the health of someone I know)
- Learn more about a medicine, test or treatment (e.g. safety, side effects or interactions)
- Find out if I or someone I know should see a doctor
- Understand medical terms
- Interpret results from blood tests or imaging
- Other (please tell us: ___________)

How much do you trust what ChatGPT says?

| Extremely | Quite a bit | Somewhat | A little bit | Not at all |
| --- | --- | --- | --- | --- |

**[Randomised to intervention group]**

[Image group] In the next section we want you to look at some information about using ChatGPT to answer your health questions. Please look at all 7 images below. There is a timer to allow you to take a closer look at the images. If you finish before the Next button appears, please have another look.

[animation group] In the next section we want you to look at some information about using ChatGPT to answer your health questions. Please watch the whole video. Once you have watched the video the Next button will appear and you can continue the survey.

[control group] In the next section we want you to look at some information about healthy eating. There is a timer to allow you to take a closer look at the image. If you finish before the Next button appears, please have another look.

**[post-intervention section]**

How much do you trust what ChatGPT says?

| Extremely | Quite a bit | Somewhat | A little bit | Not at all |
| --- | --- | --- | --- | --- |

[ChatGPT knowledge]

Please tell us which statements you agree with:

1. It’s safe to use ChatGPT in an emergency if you really need to [Yes/**no**]
2. ChatGPT can be useful for answering general health questions [**Yes**/no]
3. If ChatGPT is confident when it tells you how to interpret your blood test results, its answer is more likely to be correct [Yes/**no**]
4. You can trust ChatGPT’s answer if it references journal articles that report on clinical trials [Yes/**no**]
5. *It is best to avoid asking ChatGPT for information about a medication’s possible side effects. [Yes/no]
6. ChatGPT knows which treatment option is best for you [Yes/**No**]

*On reflection this item wording was unclear and is not included in the analysis.

List four tips that may help make ChatGPT’s answers to health questions more accurate*

1: _________

2: _________

3: _________

4: _________

*This variable is not analysed in this manuscript.

[intention to use ChatGPT in health-based scenarios]

Imagine that your uncle has just found out they have gout. The doctor has asked them to get some blood tests done including a test for uric acid. They also mentioned that gout can affect your kidneys. You want to help them find out more about this health condition. You’re thinking about using ChatGPT to get you started.

Which of the following questions would you ask ChatGPT ? [ definitely avoid | probably avoid | not sure | probably try | definitely try]

1. What is gout?
2. How is gout usually treated?
3. Should my uncle use corticosteroids to treat gout?
4. What does the uric acid test measure?
5. Is it OK if his uric acid test result is 8 mg/dL?
6. You want to learn more about how gout affects the kidneys. You find some information about gout in a well-known medical journal. You think about asking ChatGPT to summarise the information in simple terms.
7. Now imagine that your uncle has been feeling unwell. You're not sure what these new symptoms mean and whether he needs to see a doctor. You type the text below into ChatGPT to help you and your uncle decide:
   *For the last few weeks my uncle has had a rash near his eyes. Some parts of the rash have become itchy and have raised red lumps that are getting bigger. It is worse at night than during the day. Does he need to see a doctor about the rash?*

[Image/animation groups only] The next section asks you questions about what you thought of the ChatGPT information you saw earlier.

Show how much you agree with the following*:

[7 point scale from Strongly disagree to Strongly agree]

a. I found the information was created personally for me

b. I felt that the information was relevant to me

c. I felt that the information was designed specifically for me

*not analysed in this manuscript

[perceived effectiveness] Please show below how you felt about the information/video you just saw.

The information/video was…

[5 point scale from Strongly disagree to Strongly agree]

a. worth remembering

b. attention-grabbing

c. powerful

d. informative

e. meaningful

f. convincing

 [intentions to share on social media]

If you were to see this information/videos online, how likely would you share this information/videos to your socials?

[7 point scale from Very Unlikely to Very Likely]

[perceived usefulness]

Please rate the extent to which you agree or disagree with the following statements:

[5 point scale from Strongly disagree to Strongly agree]

1. The information from this study was new to me.
2. The information gave me useful tips for asking ChatGPT health questions
3. The information helped me think more carefully about which health questions to ask ChatGPT
4. The information helped me think more carefully about how to ask ChatGPT health questions
5. The information helped me think about how to use ChatGPT with other sources of information
6. I intend to apply the information I learnt in this study next time I want to ask ChatGPT a health question.
7. I am confident that I can use this information next time I ask ChatGPT a health question.

Do you have any comments about the information you saw? E.g. How could we improve it?

[free text responses]

Is there anything more that you want to know about using ChatGPT to answer your health questions?

[free text response]

[end of survey – once submitted participants in all groups will receive a link to the two social media interventions]
