## Supplementary Tables and Figures for "Can co-designed educational interventions help consumers think critically about asking ChatGPT health questions? Results from a randomised-controlled trial"

### Appendix B: Supplementary tables and figures

**Table S1. Summary of co-design activities**

| **Meeting** | **Activities / aims** |
| --- | --- |
| **1: Establish project scope** | The researchers presented an overview of ChatGPT and findings from their previous ChatGPT study.^1^ The panel then completed a brainstorming activity to gather examples of using ChatGPT for health questions and confidence asking these questions. This was followed by a discussion about comparing output from Google snippets and ChatGPT. Members had opportunity to raise further questions about using ChatGPT for health questions. Finally, the panel members and researchers collaboratively described the project scope including intended audience and format of trial resources. |
| **2: Refine key educational messages** | Key educational messages were drafted and presented to the panel. Key topics are shown in Table 2. The panel gave feedback on the key messages and associated content, including the tone/language, caveats they felt were important, and any content that was unclear |
| **3: Present content and gather feedback** | Key messages were converted into a set of social media images and a script for an animation. Panel members gave feedback on these. |
| **4: Revise content** | Panel members viewed revised images and a draft animation, as well as feedback on drafted outcome measures and survey design. Panel members discussed ways to recruit participants (social media). On piloting, further changes were discussed to refine the outcome measures further and identify a more feasible recruitment pathway. |
| **5: Present and discuss findings** | The researchers presented preliminary findings to the panel and elicited a discussion about the findings. Panel members had opportunity to view the draft paper and provide feedback. Panel members discussed potential ways to disseminate the results. Panel members were also asked to reflect on the study processes and how they were engaged as consumers. |

**Table S2. Key messages**

| **Component** | **Key messages** |
| --- | --- |
| **Basic ChatGPT concepts ^2^** | - ChatGPT doesn’t evaluate or synthesise evidence. - It can be hard to know where ChatGPT gets its information from. Even when it gives sources, these can be fake or incorrect. - It is risky to use ChatGPT as a single source of truth. - ChatGPT’s confidence is not related to the accuracy of its output. - Don’t use ChatGPT in an emergency |
| **Understanding potential risks of using ChatGPT for health** ^3-8^ | - General health questions are less risky e.g. understanding a medical term, a health condition, treatment options, or summarising health information from a reliable source. - Answers to general questions may be incorrect, missing key points, or lack important personal context. - Specific health questions that relate to personal health decisions/actions are more risky e.g. advice on treatment, diagnosis, and interpretation of medical tests. - In addition to the issues for general health questions, specific questions may cause harm including unnecessary stress or worry |
| **Strategies to reduce the risk associated with using ChatGPT for health** ^3-10^ | - If you want to use ChatGPT, focus on general questions and avoid asking about which treatment to try, diagnosis, or interpretation of medical tests. - Speaking with a health professional is always the safest option. - Compare ChatGPT’s advice to information from reliable sources. - Advice for prompts: (1) Provide context where possible and that you are comfortable to share e.g. gender/age/country but not personal information; (2) Understand the data that ChatGPT uses i.e. it may be old; (3) Break down prompts into steps; (4) Experiment with different prompts including role-playing |

**Table S3. ChatGPT knowledge score and trust in ChatGPT at followup, by intervention group**

|  | **Knowledge score** | | **Trust in ChatGPT (follow-up)** | |
| --- | --- | --- | --- | --- |
| **Intervention group** | **M** | **SD** | **M** | **SD** |
| Animation | 4.13 | 1.08 | 2.62 | 0.90 |
| Images | 3.71 | 1.39 | 2.85 | 0.95 |
| Control | 3.14 | 1.34 | 3.13 | 0.93 |

*Note*: Possible score range 0 to 5.

**Table S4. ChatGPT knowledge score by intervention group and individual knowledge item, number and per cent correct**

|  | **Animation** | | **Images** | | **Control** | | **Total** | |
| --- | --- | --- | --- | --- | --- | --- | --- | --- |
| **Knowledge item** | **n** | **%** | **n** | **%** | **n** | **%** | **n** | **%** |
| It’s safe to use ChatGPT in an emergency if you really need to | 172 | 90.1 | 155 | 76.4 | 113 | 57.1 | 440 | 74.3 |
| ChatGPT can be useful for answering general health questions | 177 | 92.7 | 180 | 88.7 | 171 | 86.4 | 528 | 89.2 |
| If ChatGPT is confident when it tells you how to interpret your blood test results, its answer is more likely to be correct | 149 | 78.0 | 138 | 68.0 | 107 | 54.0 | 394 | 66.6 |
| You can trust ChatGPT’s answer if it references journal articles that report on clinical trials | 118 | 61.8 | 116 | 57.1 | 81 | 40.9 | 315 | 53.2 |
| ChatGPT knows which treatment option is best for you | 172 | 90.1 | 165 | 81.3 | 149 | 75.3 | 486 | 82.1 |

**Table S5. ANOVA regression models for effect of intervention group and selected demographic variables (health literacy, digital health literacy, baseline trust in ChatGPT, gender, and age) on intentions to use ChatGPT for health questions.**

| **ChatGPT health scenario** | **Main effect: Intervention group** | | **Main effect: demographic variable** | | **Interaction effect: intervention group X demographic variable** | |
| --- | --- | --- | --- | --- | --- | --- |
|  | **F** | **p** | **F** | **p** | **F** | **p** |
| **Lower risk scenarios** |  |  |  |  |  |  |
| Health literacy | 0.12 | 0.884 | 20.92 | **<0.001** | 0.16 | 0.853 |
| Digital health literacy | 0.47 | 0.628 | 14.62 | **<0.001** | 1.61 | 0.200 |
| Trust in ChatGPT (baseline) | 0.05 | 0.955 | 6.35 | **0.012** | 2.28 | 0.103 |
| Age | 0.43 | 0.653 | 3.15 | 0.076 | 7.11 | **0.001** |
| Gender | 0.22 | 0.799 | 1.53 | 0.216 | 0.55 | 0.580 |
| **Higher risk scenarios** |  |  |  |  |  |  |
| Health literacy | 11.41 | **<0.001** | 2.62 | 0.106 | 0.64 | 0.528 |
| Digital health literacy | 10.00 | **<0.001** | 1.53 | 0.217 | 0.29 | 0.752 |
| Trust in ChatGPT (baseline) | 29.45 | **<0.001** | 27.44 | **<0.001** | 5.00 | **0.007** |
| Age group | 22.03 | **<0.001** | 0.38 | 0.539 | 2.33 | 0.098 |
| Gender | 22.56 | **<0.001** | 0.80 | 0.782 | 1.45 | 0.237 |

Health literacy was assessed using the single item screener.^11^ Digital health literacy was assessed using the eHeals.^12^ The cut-off for low was based on scores below the midpoint of the scale. The effect of age group was based on participants aged 18 to 44 years vs 45+ years. The effect of gender compared man or male and woman or female. Scores indicating trusting ChatGPT ‘Extremely’ or ‘Quite a bit’ were considered ‘high.’ All other response options (‘Not at all,’ ‘A little bit’ and ‘Somewhat’) were considered low.

**Table S6. Intention to use ChatGPT for health questions by intervention group, health literacy, digital health literacy, and baseline trust in ChatGPT***

| **ChatGPT health scenario** | **Animation** | | **Images** | | **Control** | |
| --- | --- | --- | --- | --- | --- | --- |
|  | **M** | **SD** | **M** | **SD** | **M** | **SD** |
| **Lower risk scenarios** |  |  |  |  |  |  |
| Limited/marginal health literacy | 3.59 | 0.78 | 3.62 | 0.85 | 3.56 | 0.92 |
| Adequate health literacy | 4.03 | 0.68 | 3.95 | 0.83 | 3.96 | 0.77 |
| Low digital health literacy | 3.46 | 0.69 | 3.78 | 0.89 | 3.61 | 0.99 |
| High digital health literacy | 4.01 | 0.70 | 3.92 | 0.83 | 3.95 | 0.76 |
| Low trust in ChatGPT | 3.95 | 0.69 | 3.83 | 0.84 | 3.80 | 0.81 |
| High trust in ChatGPT | 3.92 | 0.79 | 4.07 | 0.82 | 4.13 | 0.77 |
| **Higher risk scenarios** |  |  |  |  |  |  |
| Limited/marginal health literacy | 2.59 | 0.86 | 2.96 | 0.92 | 3.13 | 1.02 |
| Adequate health literacy | 2.37 | 1.05 | 2.64 | 1.11 | 3.12 | 0.99 |
| Low digital health literacy | 2.39 | 0.79 | 2.55 | 1.14 | 2.91 | 0.96 |
| High digital health literacy | 2.42 | 1.05 | 2.71 | 1.09 | 3.17 | 1.00 |
| Low trust in ChatGPT | 2.36 | 0.97 | 2.58 | 1.03 | 2.87 | 0.95 |
| High trust in ChatGPT | 2.55 | 1.13 | 2.94 | 1.20 | 3.74 | 0.82 |

*Higher scores indicate higher intention of using ChatGPT. A score of 1 refers to definitely avoid and a score of 5 refers to definitely try to use ChatGPT to answer the health question in the scenario(s). Health literacy was assessed using the single item screener.^11^ Digital health literacy was assessed using the eHeals.^12^ The cut-off for low was based on scores below the midpoint of the scale. Scores indicating trusting ChatGPT ‘Extremely’ or ‘Quite a bit’ were considered ‘high.’ All other response options (‘Not at all,’ ‘A little bit’ and ‘Somewhat’) were considered low.

**Table S7. Intention to use ChatGPT for health questions by intervention group, gender, and age group**

| **ChatGPT health scenario** | **Animation** | | **Images** | | **Control** | |
| --- | --- | --- | --- | --- | --- | --- |
|  | **M** | **SD** | **M** | **SD** | **M** | **SD** |
| **Lower risk scenarios** |  |  |  |  |  |  |
| Age group: 18 to 44 years | 4.01 | 0.71 | 3.89 | 0.78 | 3.65 | 0.90 |
| Age group: 45 years or more | 3.88 | 0.73 | 3.91 | 0.89 | 4.10 | 0.67 |
| Gender: Man or male | 3.87 | 0.66 | 3.90 | 0.78 | 3.81 | 0.86 |
| Gender: Woman or Female | 3.99 | 0.76 | 3.89 | 0.88 | 3.95 | 0.78 |
| **Higher risk scenarios** |  |  |  |  |  |  |
| Age group: 18 to 44 years | 2.53 | 1.07 | 2.62 | 1.06 | 3.00 | 0.98 |
| Age group: 45 years or more | 2.32 | 0.97 | 2.75 | 1.12 | 3.23 | 1.00 |
| Gender: Man or male | 2.39 | 0.95 | 2.82 | 1.03 | 3.07 | 0.98 |
| Gender: Woman or Female | 2.44 | 1.07 | 2.59 | 1.13 | 3.18 | 1.00 |

*Higher scores indicate higher intention of using ChatGPT. A score of 1 refers to definitely avoid and a score of 5 refers to definitely try to use ChatGPT to answer the health question in the scenario(s).

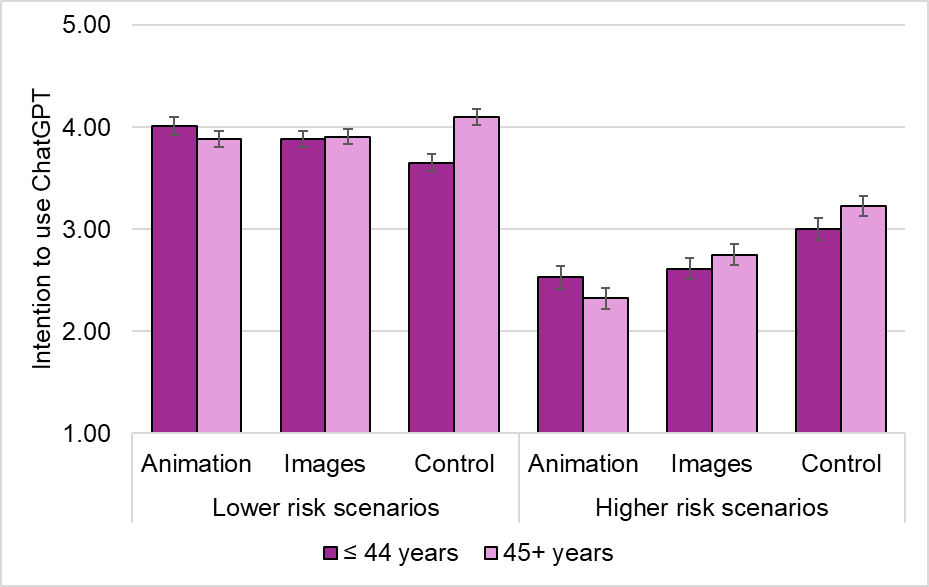

**Figure S1. Intention to use ChatGPT, by intervention group, age group and scenario type.**

*Note:* Error bars indicate ±1 SE. Higher scores indicate higher intention of using ChatGPT.

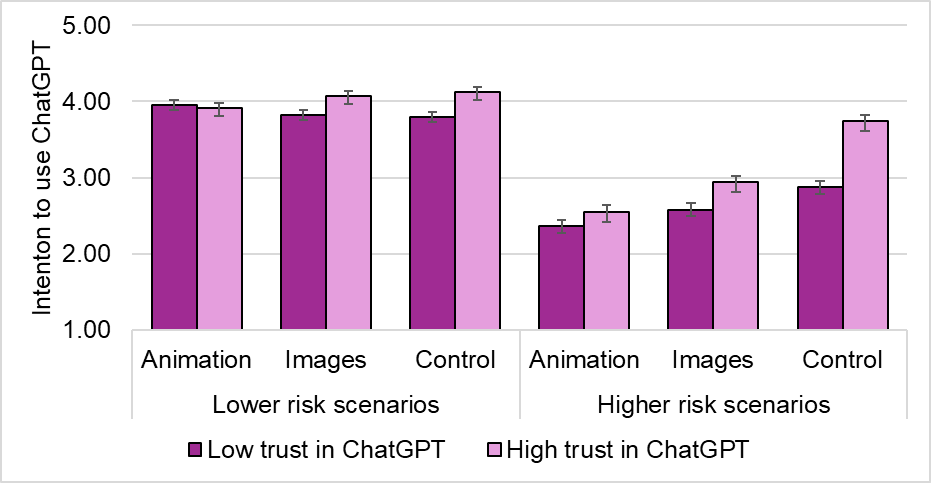

**Figure S2. Intention to use ChatGPT, by intervention group, baseline trust in ChatGPT and scenario type.**

*Note*: Error bars indicate ±1 SE. Higher scores indicate higher intention of using ChatGPT. Scores indicating trusting ChatGPT ‘Extremely’ or ‘Quite a bit’ were considered high. All other response options (‘Not at all,’ ‘A little bit’ and ‘Somewhat’) were considered low

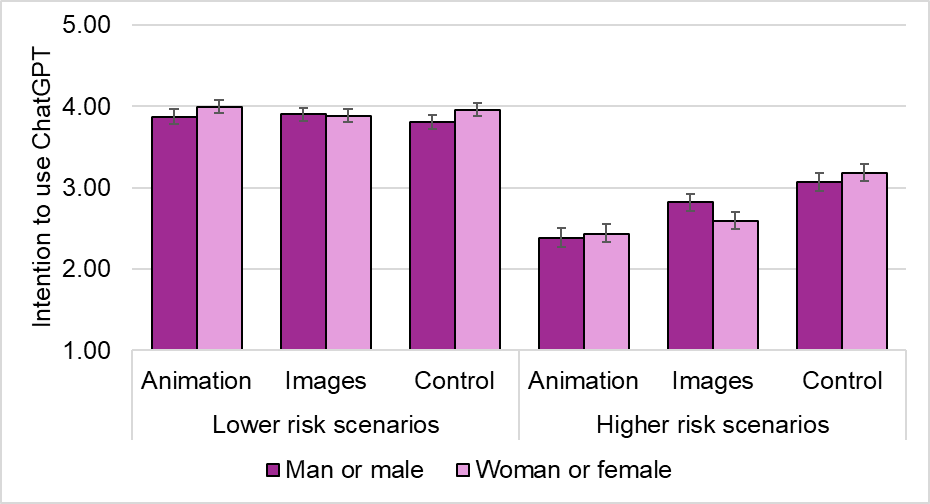

**Figure S3. Intention to use ChatGPT, by intervention group, gender and scenario type.**

*Note*: Error bars indicate ±1 SE. Higher scores indicate higher intention of using ChatGPT. The effect of gender compared man or male and woman or female.

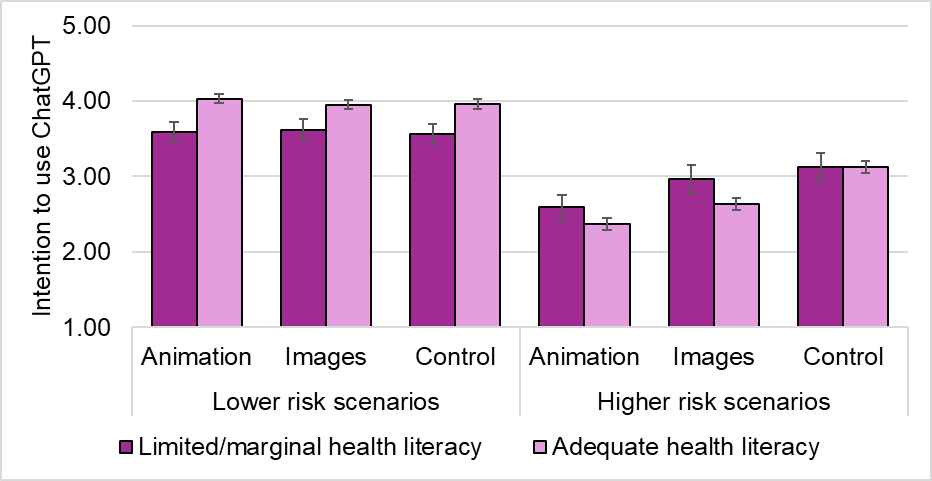

**Figure S4. Intention to use ChatGPT, by intervention group, health literacy and scenario type.**

*Note*: Error bars indicate ±1 SE. Higher scores indicate higher intention of using ChatGPT. Health literacy was assessed using the single item screener ^11^.

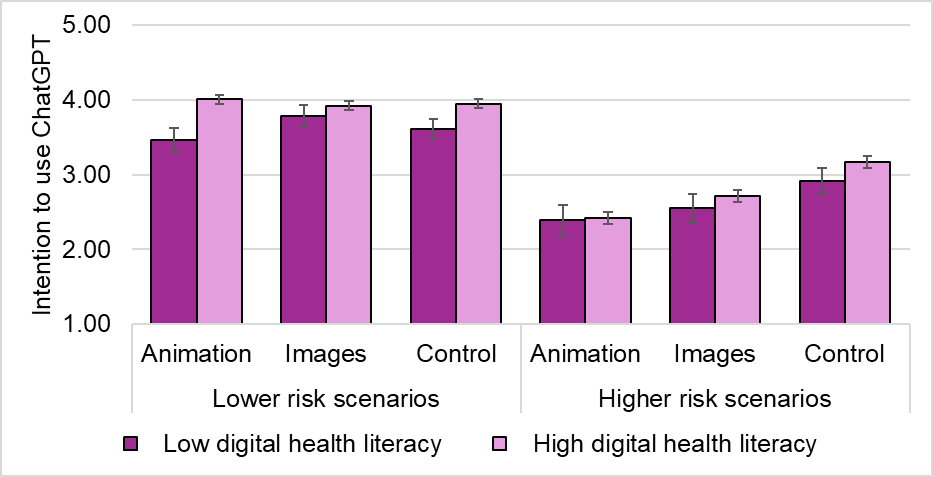

**Figure S5. Intention to use ChatGPT, by intervention group, digital health literacy and scenario type.**

*Note*: Error bars indicate ±1 SE. Higher scores indicate higher intention of using ChatGPT. Digital health literacy was assessed using the eHeals ^12^. The cut-off for low was based on scores below the midpoint of the scale.
